# Multi-Omics Integration for Identification of Prognostic Molecular Signatures for Survival Stratification in Lung Cancer

**DOI:** 10.64898/2026.02.28.26347335

**Authors:** Chayan Maitra, Vivek Das, Dibyendu B. Seal, Rajat K. De

## Abstract

Lung cancer is characterized by profound intratumoral and inter-patient heterogeneity, spanning histological subtypes, molecular landscapes, and the tumor microenvironment. While multi-omics integration is essential for capturing this complexity, leveraging these data to explicitly define survival-associated subpopulations remains a significant challenge. In this study, we developed NeuroMDAVIS-FS, an unsupervised deep learning framework designed to stratify lung cancer patients by survival risk, and identify molecular determinants underlying improved clinical outcomes. Using the CPTAC cohort, we integrated genomic (CNV), transcriptomic (RNA-seq), and proteomic profiles to extract modality-specific features. Candidate biomarkers were validated through Kaplan-Meier (KM) survival analysis and univariate Cox proportional hazards (CoxPH) regression. A final multivariate CoxPH model effectively stratified patients into high-risk and low-risk cohorts (Kaplan Meier *p*-value *<* 0.001). Notably, the integration of these molecular features with baseline clinical models significantly enhanced prognostic accuracy, improving the concordance index by 43.79% in LUAD, 31.05% in LSCC, and 23.76% across the pan-lung cancer cohort. These results demonstrate that NeuroMDAVIS-FS identifies robust, biologically relevant features that surpass traditional clinical variables in predicting patient outcomes, offering a scalable path for precision oncology.

## 1 Introduction

Lung cancer remains one of the leading causes of cancer-related mortality worldwide, with a high degree of heterogeneity at the molecular level and a complex tumour microenvironment. Despite surgical treatment, up to 30% of Stage I lung cancer patients experience recurrence within five years of diagnosis [1]. Lung cancer, specifically lung squamous cell carcinoma (LSCC) and lung adenocarcinoma (LUAD), exhibits distinct molecular profiles and clinical behaviors, contributing to their complexity. Both subtypes share some common genetic alterations, such as mutations in the *TP*53 gene, but differ significantly in mutational landscapes and histological characteristics, affecting treatment strategies and prognosis. According to the Indian Council of Medical Research–NCDIR National Cancer Registry Programme (2020), lung cancer ranks among the leading cancers in India, with tobacco-associated cancers accounting for nearly 27.1% of the overall cancer burden, and a marked rise in lung cancer incidence reported across the majority of registries ^1^. In the United States, lung cancers are currently the second leading cause of new cancer cases for both males and females ^2^. Therefore, identifying reliable biomarkers for early-stage lung cancer remains an urgent medical priority.

Biomarker prediction has long been supported by survival analysis as a cornerstone of clinical translational research, particularly in oncology, where time-to-event outcomes, such as overall survival and progression-free survival, are considered primary endpoints. Classical approaches based on the Cox proportional hazards model [2] were widely employed to identify prognostic biomarkers by associating molecular features with patient survival while accounting for censoring. As omics data are inherently high-dimensional and multicollinear, regularized extensions, such as LASSO-Cox and elastic-net Cox models, were proposed [3], enabling simultaneous feature selection and risk estimation. More recently, there was an increasing interest in deep learning-assisted survival analysis for robust biomarker discovery driven by the exponential growth in the availability of high-dimensional omics data and by recognized limitations of traditional Cox proportional hazard models in mimicking complex, nonlinear relationships. These methods are at the core of developing multivariate prognostic signatures and risk scores that outcompete single-marker analyses and support patient stratification into clinically meaningful risk groups. The first extensions of survival analysis using neural networks, for example, Cox-nnet [4], or DeepSurv [5], illustrated that deep feed-forward architectures can flexibly model nonlinear hazard functions while retaining the partial-likelihood framework of Cox regression, thereby enabling simultaneous feature selection (i.e., data-driven identification of survival-associated molecular features) and risk estimation. With advancements in large-scale molecular profiling, subsequent studies [6, 7] extended this paradigm towards the integration of multi-omics data along with feature selection embedded in survival-aware neural networks to allow simultaneous risk prediction and prioritization of biologically meaningful biomarkers across genomic, transcriptomic, and proteomic layers.

Advances in multi-omics technologies enabled researchers to integrate multiple layers of biological data along the central dogma, providing deeper insights into disease mechanisms. As these technologies evolved, data volumes expanded exponentially, making efficient analysis and interpretation of such high-dimensional data—especially when combined with clinical features, a significant challenge. Machine Learning (ML) and Deep Learning (DL) approaches became increasingly popular and mainstream for addressing these complex problems. Recent multi-omics integration methods, such as GLUE [8], scJoint [9], TotalVI [10], UMINT [11], MUON [12], and MATILDA [13], among others, addressed the issue of dimensionality by generating low-dimensional latent embeddings useful for downstream analyses. However, these DL methods, used in multi-omics data analysis, are often unable to generate biologically significant information. Researchers recently highlighted the growing relevance of biologically informed neural networks (BINNs) [14, 15] as a potential solution to this challenge [16]. Thus, feature selection and dimensionality reduction techniques play a crucial role in identifying the most informative biomarkers to aid patient prognosis, disease understanding, and future treatment strategies.

With the recent developments in computational pathology and deep learning methods, the scope of survival analysis further extended beyond the conventional covariates by incorporating prognostic information from histology images [17, 18, 19]. Beside genomic and transcriptomic biomarkers, it included histopathological biomarkers [20], thus enabling more comprehensive modeling of lung cancer, along with the others. Survival-aware risk scores were also incorporated to enable the possibility of data-driven biomarker discovery for lung squamous cell carcinoma and other subtypes. The recognition of the heterogeneity of lung cancer also underscored the need for robust survival modeling, which can enable the generalizability of lung cancer models. Survival analysis thus emerged as a tool that not only enables prediction but also bridges the gap between lung cancer, molecular biomarkers, and treatment strategies. However, despite enhanced predictive accuracy, challenges persist in terms of model interpretability, cohort generalizability, and biological validation of predicted biomarkers, underscoring the need for integrative strategies that combine survival modeling with pathway-level and functional analyses.

In this study, we developed NeuroMDAVIS-FS, a feature selection method to analyse two subtypes of lung cancer, viz., LSCC and LUAD. The model is built upon NeuroMDAVIS [21] that comprised a latent layer, a shared hidden layer, and a modality-specific hidden layer, which projected multi-omics data into a common latent space while minimizing reconstruction loss. NeuroMDAVIS-FS examines the weights and biases of the trained NeuroMDAVIS model to rank features according to their importance. The goal of this work was to identify clinically relevant features contributing to lung cancer progression.

To demonstrate the effectiveness of NeuroMDAVIS-FS, we performed a series of experiments using lung cancer datasets from the Clinical Proteomic Tumor Analysis Consortium (CPTAC) [22], having Copy Number Variation (CNV), RNA expression, and protein expression data. First, we trained NeuroMDAVIS to obtain a low-dimensional latent representation of the multi-omics data. We then applied NeuroMDAVIS-FS to identify the most significant features in each omics layer. Later, the selected features were assessed using survival analysis through KM statistics and univariate CoxPH, segregating patients into low expression and high expression groups. Thereupon, a multivariate CoxPH model was fitted to stratify patients into high-risk and low-risk groups, which displayed significant differences in survival. Eventually a baseline model was built using clinical features, and compared against the features selected by NeuroMDAVIS-FS. Adding the selected features to the baseline model improved the concordance index by a significant margin. Thus, this study offered an advanced framework for identifying clinically-relevant features in lung cancer. The overall framework is represented as Figure 1.

**Figure 1:**
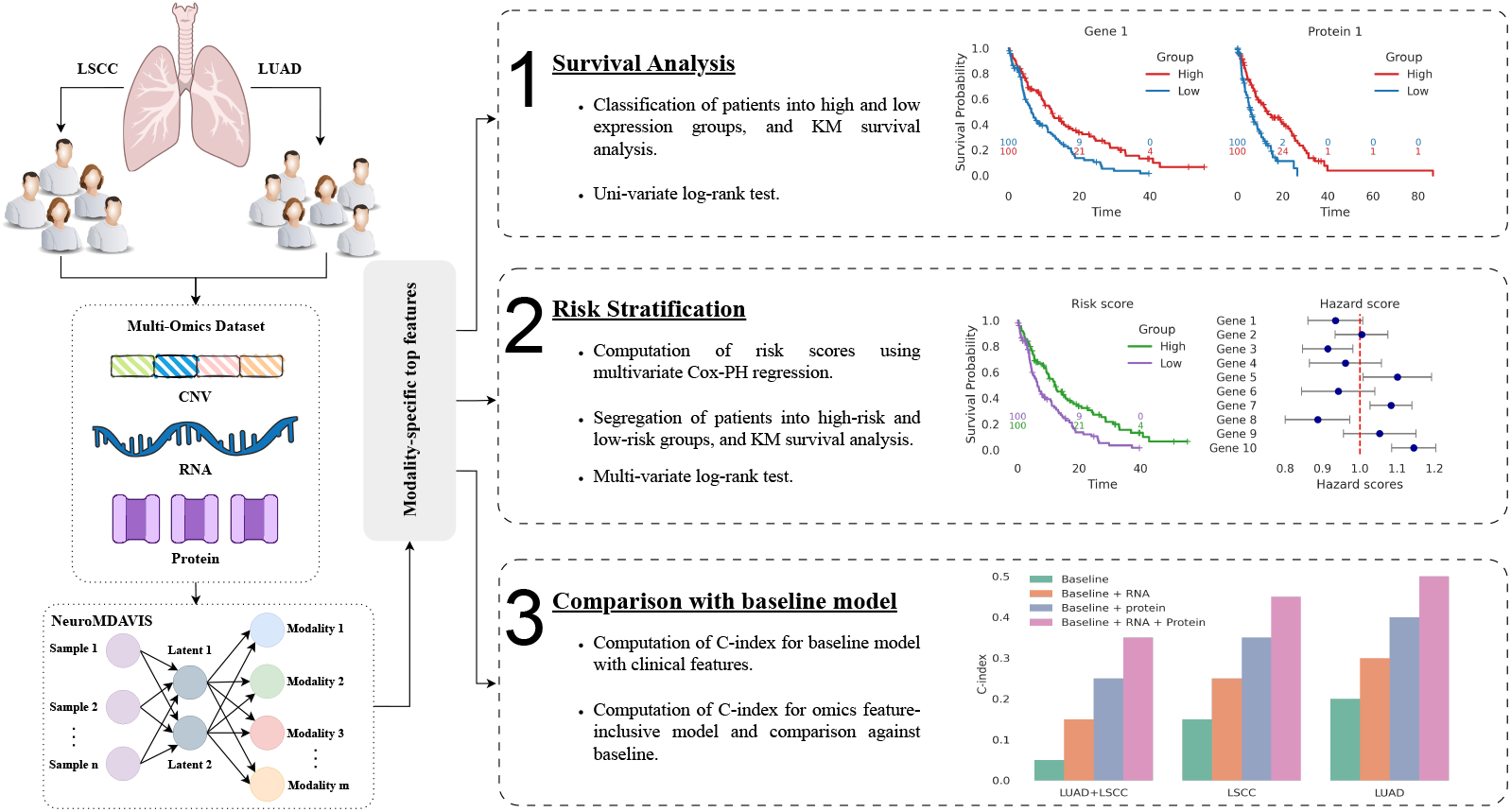
Graphical overview of the multi-modal survival analysis framework using modality-specific top features. (1) Survival analysis was performed by stratifying patients into high and low-expression groups based on individual modality-specific features, followed by Kaplan–Meier (KM) survival estimation and univariate log-rank testing. (2) Risk stratification was conducted using multivariate Cox proportional hazards (CoxPH) regression to compute patient-specific risk scores, enabling separation into high and low-risk groups. (3) Model performance was evaluated by comparing the concordance index (C-index) of a baseline clinical model with omics-augmented models across LSCC, LUAD, and the combined cohort, demonstrating improved prognostic performance with multi-modal integration.

## 2 Methodology

In this section, we discuss the datasets used in the experiments and the proposed methodology for selecting important features from multi-omics data. We then outline the experimental design used to evaluate the effectiveness of the selected features through survival analysis, risk stratification, and comparison with clinical features.

### 2.1 Data acquisition and pre-processing

Proteogenomic datasets for Lung Squamous Cell Carcinoma (LSCC) and Lung Adenocarcinoma (LUAD) were obtained from CPTAC data portal. Data cohorts include clinical information, genomics, transcriptomics, and proteomics profiles for 205 patients (103 LSCC, and 102 LUAD). For each cancer type, raw data files were downloaded and parsed using the CPTAC Python package (version 1.5.14) [23]. Samples with missing clinical annotations or low-quality omics data were excluded. Omics data were log2-transformed, normalized, and preprocessed using Python 3. The distribution of demographic and clinical features is shown in Figure 2.

**Figure 2:**
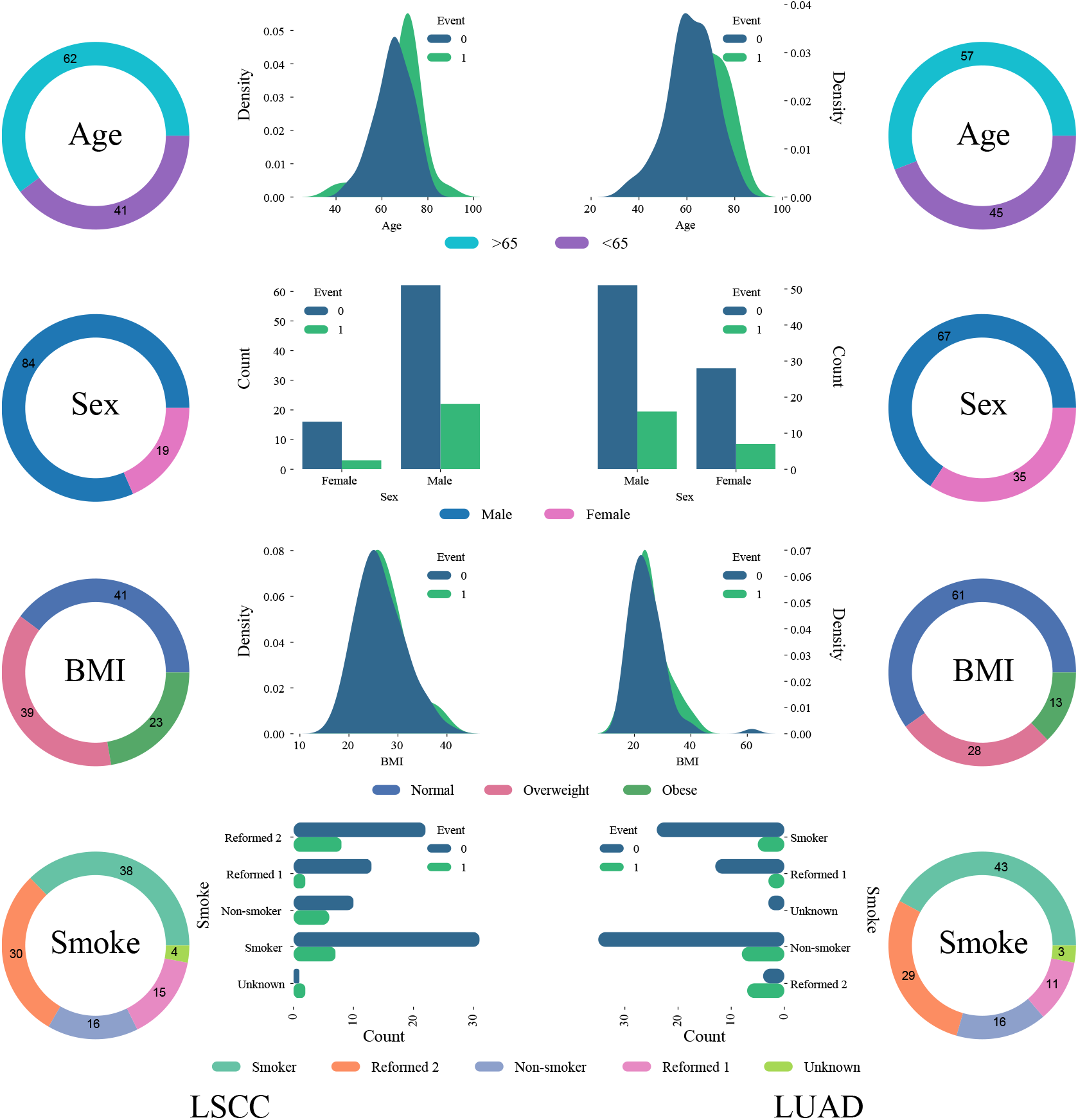
Event-wise distribution of demographic and clinical variables. Kernel density plots show differences in Age and BMI between Event = 0 (censored) and Event = 1 (death), while bar plots depict event-stratified counts for Sex and Smoking status. Donut charts summarize the overall categorical composition for each variable (age groups, BMI classes, sex, and smoking categories). The left and right panels show the details of LSCC and LUAD cohorts, respectively. ‘Censored’ refers to an event that has not yet occurred within the study’s timeframe.

### 2.2 NeuroMDAVIS-FS

NeuroMDAVIS-FS is a feature selection model built upon the NeuroMDAVIS framework, specifically designed to identify the most informative features in multi-modal datasets. Here we considered multi-omics data to demonstrate its effectiveness. NeuroMDAVIS itself, is an unsupervised neural network framework designed for multi-omics data visualization, where the objective is to reconstruct the input omics modalities [21]. We demonstrated earlier that the feature embedding generated by NeuroMDAVIS, solely obtained through reconstruction, was highly informative. Based on the assumption that highly informative features are reconstructed with greater precision than non-informative ones, NeuroMDAVIS-FS was developed. Additionally, we know that feature variance strongly influences the reconstruction process, i.e., features with little to no variability are reconstructed more accurately since their values remain consistently aligned across samples. However, these features may not be informative enough. Hence, the objective function was designed to reinforce the claim that a feature is said to be informative only if it demonstrates sufficient variability and is also effectively reconstructed by NeuroMDAVIS after training.

In order to formalize this notion, we introduced a scoring scheme as explained below.

Let **D** be a collection of datasets each for one of *m* different omics modalities.

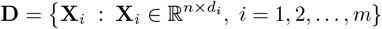

The *i*^*th*^ omics modality **X**_*i*_ contains *n* samples and is characterized by *d*_*i*_ features, that is,

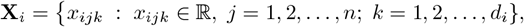

where *x*_*ijk*_ denotes *k*^*th*^ feature of *j*^*th*^ sample from the *i*^*th*^ modality. The *input layer* of NeuroMDAVIS consists of *n* neurons which take an identity matrix as input and generate lossy reconstructions 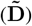 of each omics modality, where

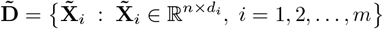

Let us assume a few more notations as follows:

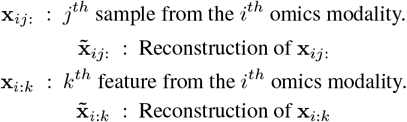

Initially, the reconstructed features are compared with the original ones for all the omics modalities with a Kullback-Leibler Divergence (KLD) loss which is given by

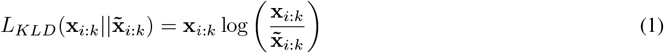

A close reconstruction of the original feature results in the loss approaching zero. Conversely, as the deviation between the reconstructed and original feature increases, the loss also increases. Formally,

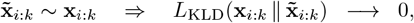

Let 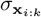 represent the standard deviation of the *k*^th^ feature in the *i*^th^ omics modality. It is well-known that a feature with little to no variability can be reconstructed easily, since these values are close to a constant direction, which makes the feature less informative for downstream tasks. On the other hand, features with higher variance are more likely to capture meaningful biological or statistical differences across samples.

Therefore, an informative feature should satisfy two criteria simultaneously:

- It should be reconstructed well by NeuroMDAVIS, indicated by a smaller 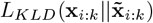, and
- It should exhibit sufficient variability, reflected by a larger 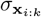.

Based on these two criteria, we define a feature score that balances the trade-off between reconstruction quality and variability. Mathematically, the combined score function for *k*^*th*^ feature in the *i*^*th*^ modality is defined as

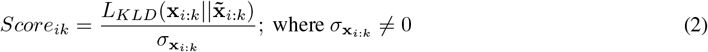

Equation (2) suggests that the lower the combined score, the better is the importance of the feature.

Finally, the set of top *t* informative features (*IF*_*i*_) from each omics modality was selected based on these scores. That is,

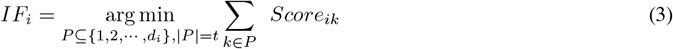

These top *t* features were then further utilized for several downstream analyses.

### 2.3 Experimental Design

In order to evaluate the prognostic significance of molecular features in lung squamous cell carcinoma (LSCC) and lung adenocarcinoma (LUAD), we developed a multi-step experimental pipeline that integrated feature selection, survival analysis, risk stratification, and model validation. High-dimensional omics data, including CNV, RNA, and protein expression values, were utilized for selecting top informative features using NeuroMDAVIS-FS. These top *t* features were then further utilized for these downstream analyses.

#### Survival Analysis

In this study, KM survival analysis, using time-to-event data, was considered to estimate survival probabilities, model risk rates and to identify prognostic factors. The selected set of top features including the ones chosen from each modality, was used to predict patient survival. Patients were then stratified into high and low-expression groups based on the median value of each feature, and log-rank tests were conducted to assess statistical significance in survival differences.

#### Risk Stratification

Risk stratification is the process of classifying individuals into different risk groups (e.g., high-risk vs. low-risk) based on their predicted likelihood of experiencing an event (e.g., death). We applied CoxPH, a semi-parametric regression model on the top-selected features across omics modalities. It helped us to find the relationship between covariates (e.g., clinical features) and the time until an event occurs. Subsequently, a risk score for each patient was computed based on the coefficients of the multivariate CoxPH model. The patients were then categorized into high-risk and low-risk groups using median risk scores, and survival outcomes between these two groups were studied using KM curves. The hazard ratios and the hazard indices were then critically assessed to ensure reliability of the CoxPH model. To account for potential covariate interactions and validate robustness of findings across several experiments, we evaluated statistical significance of survival differences using both univariate and multivariate log-rank tests.

#### Comparison with baseline model

In order to quantify the incremental prognostic value of the omics-derived signatures, we established a baseline CoxPH model incorporating standard clinical covariates, including age, sex, BMI, and smoking history. The predictive accuracy of this baseline was then compared against the multi-omics integrated model by evaluating the improvement in the concordance index (C-index).

## 3 Results

Initially, three distinct modalities of omics data, namely CNV, RNA, and protein expressions, were fed as input to NeuroMDAVIS which integrated them to generate a low-dimensional latent embedding. This latent representation effectively condensed the complex relationships among different omics layers while preserving essential information. Thereafter, to gain deeper biological insights and improve interpretability, NeuroMDAVIS-FS (Feature Selection module) was employed. Instead of relying solely on the latent embeddings, NeuroMDAVIS-FS analysed the weights and biases of the trained NeuroMDAVIS model to identify the most informative features from the original omics data. These selected features were expected to play a crucial role in characterizing the molecular features under study. In order to assess the impact of the selected features on patient prognosis, KM survival analysis was carried out. The KM estimator was utilized to evaluate the effectiveness of the selected features in distinguishing different survival groups. Further analysis involved risk stratification using CoxPH regression and comparison against baseline model built using clinical features only.

### 3.1 Feature Selection

To highlight cancer-specific top features, we considered three scenarios. First, features were selected from the combined dataset containing both cancer types, as presented in Table 1. Next, we examined each cancer subtype individually, with the top features for LSCC (Supplementary Table S1) and those for LUAD (Supplementary Table S2). The features ranked by NeuroMDAVIS-FS were found to be highly informative since they direct towards major biochemical pathways associated with cancer in general.

**Table 1:** Top 15 molecular features selected by NeuroMDAVIS-FS from each of the omics modalities, viz., Protein, RNA, and CNV. Here both LSCC and LUAD samples were utilized to train NeuroMDAVIS.

| Rank | Protein | RNA | CNV |
| --- | --- | --- | --- |
| 1 | SLC19A3 | CDY2B | LIMD1 |
| 2 | TAL1 | DAZ4 | CXCR6 |
| 3 | KRT13 | DCAF4L2 | SLC6A20 |
| 4 | WSB2 | XKR4 | CCR9 |
| 5 | GMCL1 | PIWIL2 | CCR1 |
| 6 | SULT1C4 | PAGE2 | CCR3 |
| 7 | C22orf39 | PAGE5 | LTF |
| 8 | MCTS2P | PNMA6E | FYCO1 |
| 9 | FBXL7 | PAGE2B | CCRL2 |
| 10 | NEK11 | SLC5A5 | XCR1 |
| 11 | KRT6A | HSD17B13 | LZTFL1 |
| 12 | PPFIA2 | GP2 | CCR2 |
| 13 | SMAD6 | OBP2B | SACM1L |
| 14 | EFCAB1 | SLC28A2 | LARS2 |
| 15 | ADPRM | PPP4R3C | CCR5 |

### 3.2 Survival Analysis

The KM survival plots (Figure 3) illustrate the prognostic value of the top-selected features (genes, CNVs, or proteins) by comparing patients with high and low expression levels. This high-low split has been considered based on the median expression value of that feature.

**Figure 3:**
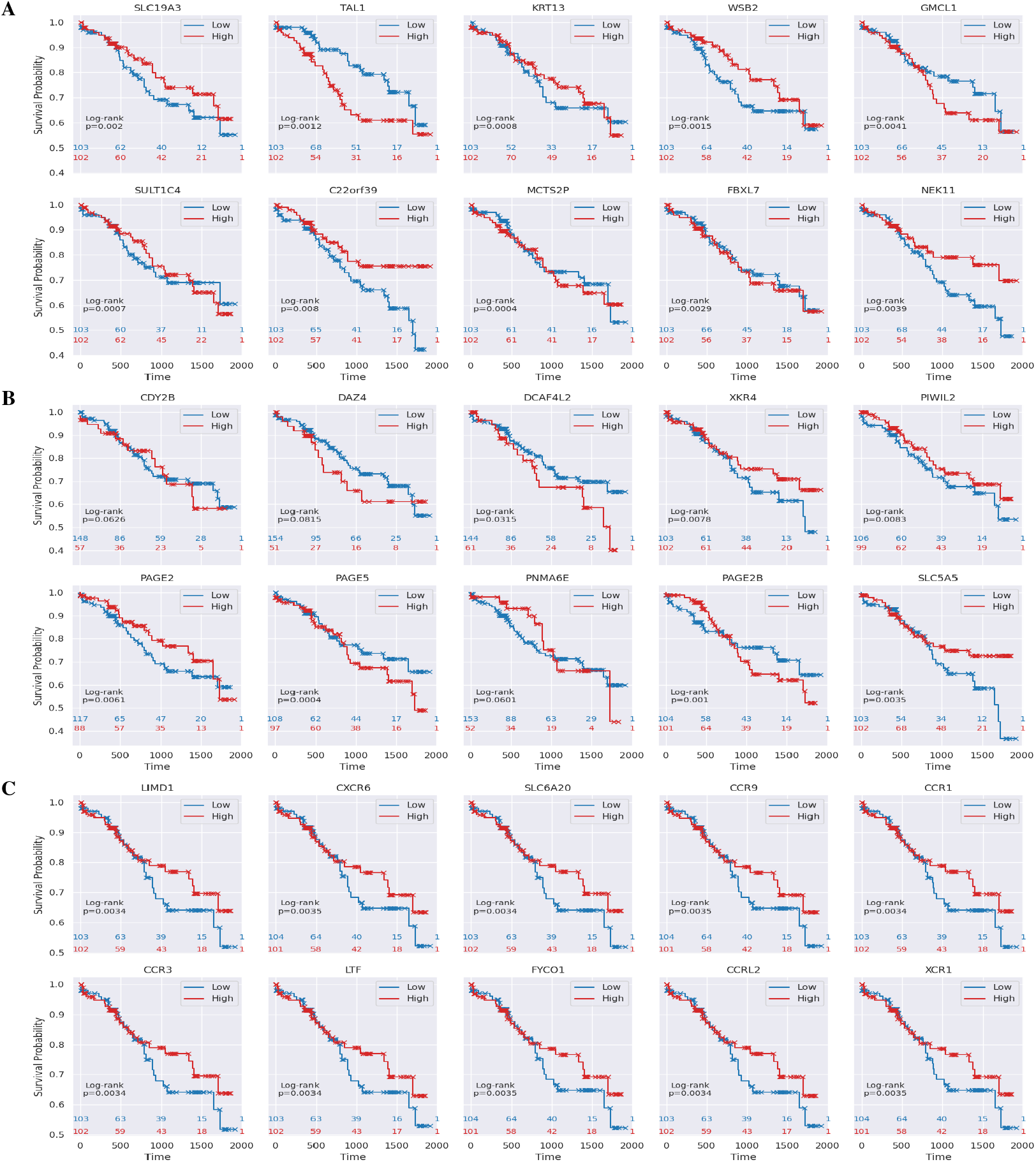
Kaplan-Meier curves based on high and low-expression values of each of the top features selected by NeuroMDAVIS-FS from (A) Protein, (B) RNA, and (C) CNV omics modalities, while applied on both the LSCC and LUAD cohort together. Patients stratified into high and low-expression groups based on the median value of each feature. Log-rank tests were conducted to assess statistical significance in survival differences.

First, as shown in Figure 3(A), some of the top ranked proteins, such as *SLC*19*A*3, *TAL*1, *KRT*13, *WSB*2 and *GMCL*1, showed significant survival differences between the two groups, with log-rank *p*-value consistently below 0.01. Notably, higher expression levels of these proteins are generally associated with either reduced or improved survival probability, suggesting their potential role in disease progression.

Second, the top features from RNA showed mixed trends in survival, as shown in Figure 3(B). While genes such as *DCAF* 4*L*2, *XKR*4, *PIWIL*2, *PAGE*2, *PAGE*5, *PAGE*2*B*, and *SLC*5*A*5 demonstrated significant differences in survival (*p*-value *<* 0.05), others like *CDY* 2*B, DAZ*4, and *PNMA*6*E* did not show significant associations (*p*-value *>* 0.05).

Finally, the top CNVs also followed a similar trend (Figure 3(C)), focusing on immune-related and transporter genes, like *LIMD*1, *CXCR*6, *SLC*6*A*20, *CCR*9, *CCR*1, *CCR*3, *LTF, FY CO*1, among others. Here too, most of the comparisons yielded statistically significant results, with log-rank *p*-values around 0.003. In these cases, patients stratified into the high-expression group often displayed survival advantages compared to the low-expression group, reinforcing the potential importance of these genes in influencing clinical outcomes, possibly through immune regulation or cellular transport pathways.

Overall, we observed that certain genes (*TAL*1, *GMCL*1, *PAGE*5) were linked to patient survival, with high expression values, predicting worse outcomes whereas others, like *SLC*19*A*3, *LIMD*1 and *CXCR*6, predicted better outcome.

The above experiment was conducted considering all samples from both LSCC and LUAD cohorts. The top molecular features from each cohort were analysed individually for predicting survival (Supplementary Figure S1 and S3).

### 3.3 Risk Stratification

In order to evaluate the prognostic potential of the selected RNA features, a multivariate CoxPH model was employed. This model estimated the hazard function by relating gene expression levels to patient survival times. Each feature contributed a regression coefficient that reflects its impact on the risk of an event. By combining these weighted contributions across all selected features, the CoxPH model generated a composite risk score for each patient, which served as the basis for stratifying individuals into high and low-risk groups. KM analysis revealed a strong separation between high and low-risk groups (Figures 4A and 4B). Patients classified as high-risk group demonstrated severely reduced survival probabilities over time, whereas those in the low-risk group consistently exhibited more favorable outcomes. This observation emphasized the prognostic value of the selected features for predicting patient survival and guiding risk-based clinical decision-making.

**Figure 4:**
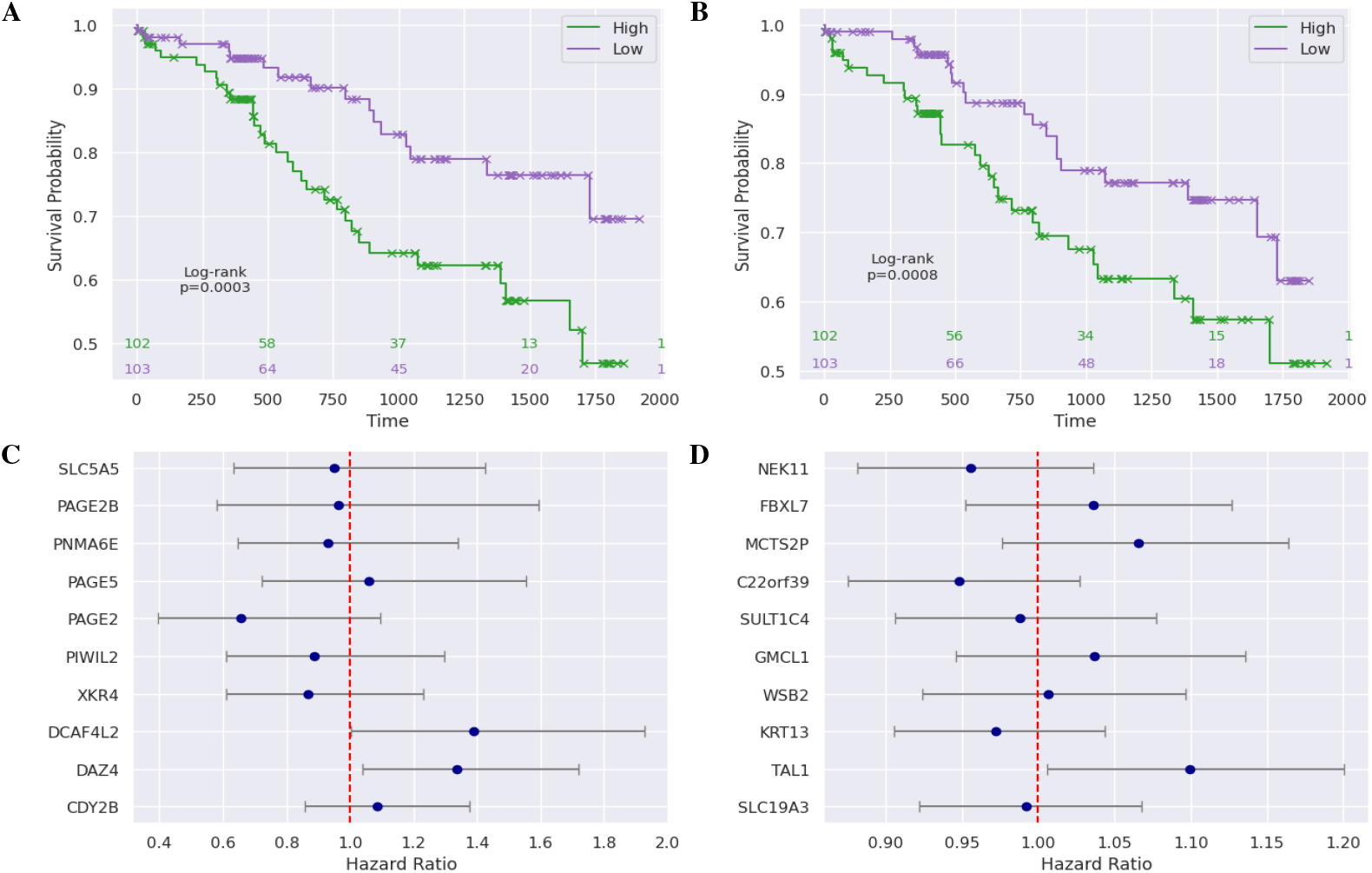
Kaplan-Meier curve based on high and low risk scores obtained from a CoxPH regression applied using (A) top RNAs and (B) top proteins, selected by NeuroMDAVIS-FS. Hazard scores of (C) top RNAs and (D) top proteins, selected by NeuroMDAVIS-FS. These results were obtained utilizing both the LSCC and LUAD cohort together.

The hazard ratio (HR) analysis of these top RNA features (Figure 4C) further supported this finding. Several genes, including *CDY* 2*B, DAZ*4, and *DCAF* 4*L*2, displayed HR values above 1.0, suggesting an association with increased risk, while others, such as *XKR*4 and *PIWIL*2, had HRs slightly below 1.0, uncorrelated with risk. The majority of RNA features demonstrated wide confidence intervals, reflecting some variability, but the overall trend indicates that these features can distinguish between higher and lower risk of poor survival outcomes.

Similarly, when evaluating the top protein features (Figure 4D), high-risk patients again exhibited a steeper decline in survival compared to low-risk patients. The hazard ratio plot showed that most of the protein features clustered close to 1.0, with *TAL*1, *FBXL*7, and *MCTS*2*P* slightly above 1.0, indicating potential risk associations, while *NEK*11 and *C*22*orf* 39 had HR values below 1.0.

These findings together demonstrated that both RNA and protein features selected by NeuroMDAVIS-FS are potentially good survival predictors. The combination of Kaplan–Meier survival analysis and hazard ratio evaluation provided strong evidence of their prognostic power, supporting their potential use for patient stratification and biomarker design. Moreover, the effect of top molecular features, obtained from individual analysis of LSCC and LUAD, also showed a significant effect on patient survival (Supplementary Figures S2 and S4 respectively).

### 3.4 Comparison with baseline

Figure 5 illustrates the performance of different survival prediction models across three groups of the dataset. These groups include both LSCC and LUAD combined, as well as LSCC and LUAD individually. Performance was measured using the concordance index (C-index). The baseline model, which incorporated only clinical variables (age, sex, BMI, and smoking history), consistently showed the lowest predictive performance across all three groups, with C-index values around 0.62. This made us belief that while clinical features offer some predictive power, they are insufficient for accurately stratifying patient outcomes on their own.

**Figure 5:**
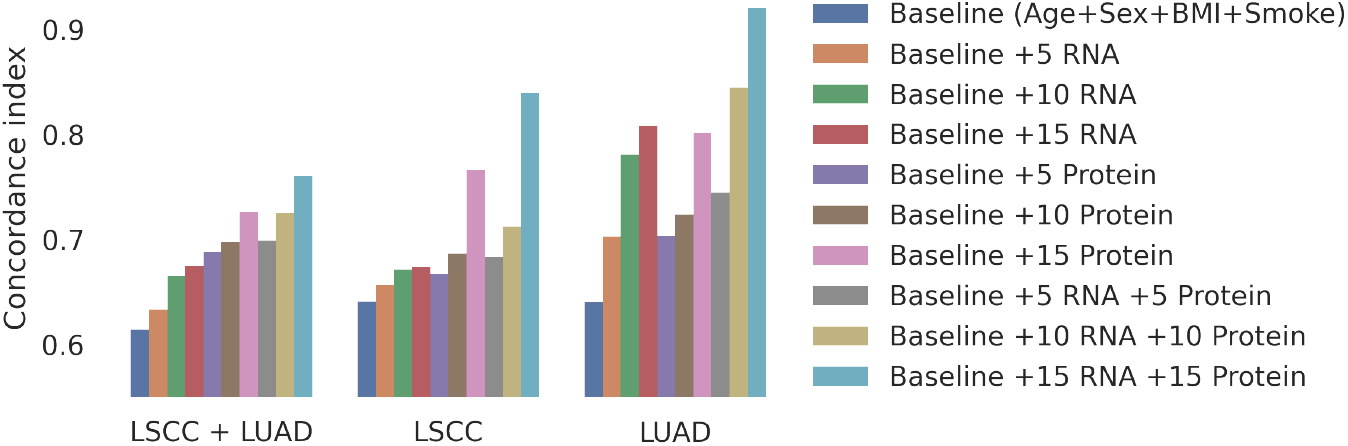
Comparison of predictive performance, measured by the concordance index (C-index), across different feature configurations and cohorts. Results are shown for a baseline clinical model comprising age, sex, BMI, and smoking status, as well as for models incrementally augmented with RNA features, protein features, and their joint combinations. Evaluations are reported for the combined LSCC-LUAD cohort, and for the individual LSCC and LUAD cohorts.

Enhancing the baseline model with RNA features improved the C-index in a stepwise manner. Adding the top 5 RNA features resulted in a modest improvement, while incorporating the top 10 and 15 RNA features led to progressively higher concordance indices. A similar pattern was observed with protein features, although the improvement appeared slightly more moderate compared to RNA features. Notably, in the LUAD group, RNA features alone substantially boosted the model’s predictive ability, with the C-index value surpassing 0.8 when 15 features were included. This highlighted the strong prognostic relevance of RNA features in this cancer subtype.

The combined model integrating both RNA and protein features alongside clinical data, achieved the highest predictive performance across all groups. In particular, for LUAD, the inclusion of 15 combined molecular features resulted in a C-index above 0.9, demonstrating excellent survival prediction accuracy. This trend indicated that integrating multi-omics features on top of clinical features, provided complementary information that significantly enhanced the model performance. In contrast, LSCC and the combined LSCC-LUAD group showed more moderate improvements, suggesting potential differences in molecular heterogeneity between these cancer types. Overall, these results emphasized the value of incorporating molecular biomarkers, especially in combination, to improve survival prediction beyond traditional clinical factors.

## 4 Discussion

The identification of these features through the NeuroMDAVIS-FS framework underscores the model’s ability to pinpoint biologically significant markers from high-dimensional multi-omics data. By integrating genomic (CNV), transcriptomic (RNA), and proteomic (protein) data, the model captured a holistic view of the tumor landscape that traditional clinical models overlook. In this study, the clinical utility of the NeuroMDAVIS-FS framework was validated through its robust ability to stratify patient cohorts into distinct risk groups. By employing a multivariate Cox proportional hazards (CoxPH) model, we successfully categorized patients into high and low-risk subpopulations characterized by significant survival disparities (Kaplan Meier *p*-value *<* 0.001). Central to the value of this deep learning-based approach is its additive prognostic power; when the identified multi-omics features were integrated with a clinical baseline model, we observed a substantial increase in the concordance index - an improvement by 43.79% in LUAD, 31.05% in LSCC, and 23.76% across the combined lung cancer cohort. These results suggest that the features selected by our model captured critical molecular drivers of mortality, which are not accounted for by standard clinical variables, highlighting the limitations of relying solely on traditional staging and the necessity of multi-omics integration in precision oncology. Interestingly, we also observed that some of the molecular hits, obtained by the model, are established tumor subtype-specific drivers, tumor suppressors, associated with metastasis, immune microenvironment and therapeutic resistance, while others can be considered novel or emerging. These hits will need future orthogonal validation studies to establish their mechanistic roles as biomarker or drug targets given their strong association with prognosis in present study. Further investigations were performed to assess the functional and clinical associations of the top molecular features obtained from each analysis, as shown in Supplementary Table S3.

### 4.1 Subtype-Specific Drivers and Tumor Suppressors

In the LUAD and the combined analysis, the selection of *LIMD*1 and *V IPR*1 in the CNV modality aligns with their established roles as critical tumor suppressors [24, 25]. *LIMD*1, located at the 3*p*21.3 locus, is a well-documented suppressor whose loss is a hallmark of lung adenocarcinoma progression [25]. Similarly, the model identified *PTPRG* (Protein tyrosine phosphatase gamma) in LSCC, a known tumor suppressor on 3*p*14.2, frequently inactivated in squamous cell carcinomas via loss of heterozygosity (LOH) [26]. These findings demonstrate that the model effectively prioritized genomic regions with high functional impact on survival.

### 4.2 Metastasis and the Immune Microenvironment

The model’s ability to select chemokine receptors, including *CCR*9, *CCR*1, *CCR*3, and *CXCR*6 from combined analysis, suggests that it captured essential components of the tumor microenvironment (TME) [27, 28, 29, 30]. *CCR*9 is a known driver of *PI*3*K*/Akt-mediated metastasis [27], while *CXCR*6 and LTF (Lactotransferrin) have been linked to immunotherapy response and immune cell infiltration [30, 31]. The identification of *ACKR*2 from the LUAD analysis, an atypical chemokine receptor that scavenges inflammatory ligands to regulate the TME, further supports the model’s ability to identify factors governing the interface between the tumor and the host immune system [32].

### 4.3 Therapeutic Resistance and Prognostic Gain

Beyond tumor biology, NeuroMDAVIS-FS identified markers of therapeutic resistance, such as *ABCC*2 (*MRP* 2) in the LUAD analysis [33]. Overexpression of this multi-drug resistance transporter is a verified predictor of cisplatin resistance in NSCLC patients, offering a biological explanation for the poor prognosis in certain identified clusters. Many of these genes or proteins, such as the chemokine receptors *CCR*1 and *CCR*3, are currently being explored as therapeutic targets [28, 29]. This suggests that the high-risk subpopulations identified by NeuroMDAVIS-FS could potentially be stratified for targeted chemokine receptor inhibitors or personalized immunotherapy regimens in future clinical trials.

## 5 Conclusion

This study demonstrates the utility and extension of deep learning-based framework for multi-omics integration for feature selection using the newly developed model, called NeuroMDAVIS-FS, which is designed to decode the molecular complexities of lung cancer. By leveraging synchronized encoding and modality-specific decoding layers, the model maps disparate data streams, including copy number variations, transcriptomes, and proteomes, into a common latent projection that preserves critical biological signals.

This approach aligns with the emerging paradigm of Biologically Informed Neural Networks (BINNs); rather than treating the tumor as a black box. NeuroMDAVIS-FS bridges the gap between raw high-dimensional data and biological interpretability. Validation using the CPTAC lung cancer dataset confirms that the framework effectively prioritizes high-impact features, such as *LIMD*1, *ABCC*2, and *CCR*9, which are deeply rooted in lung cancer pathogenesis, therapeutic resistance, and metastatic potential. By focusing on these biologically significant nodes, the model provides a transparent mechanism for precision oncology, identifying ‘vulnerability hubs’ that enable improved patient stratification compared with traditional clinical models. This is evidenced by a concordance index improvement of up to 31.05% in LSCC and 43.79% in LUAD. While NeuroMDAVIS-FS effectively captured key ‘vulnerability hubs’, the current architecture is yet to embed explicitly higher-order biological priors, such as pathway hierarchies or protein-protein interaction networks, which could further refine the model’s accuracy and biological interpretability in a precision oncology context.

Future work will focus on expanding this framework to larger and more diverse cohorts, validating the findings across multiple datasets to ensure robustness and generalizability. Such iterations will focus on explicitly embedding biological priors, such as pathway hierarchies and protein-protein interaction networks, directly into the architecture to further refine the model’s accuracy. Additionally, incorporating attention mechanisms or graph-based learning may further enhance the interpretability and accuracy of multi-omics integration.

By transforming complex multi-omics landscapes into actionable prognostic insights, this framework provides a scalable foundation for precision medicine. It enables the discovery of robust biomarkers that may guide targeted therapy development and the design of personalized immunotherapy regimens. Ultimately, NeuroMDAVIS-FS offers an advanced, interpretable deep learning paradigm that paves the way for more precise prognostic tools and improved clinical outcomes in lung cancer research.

## Data and Code availability

The datasets and codes used in this study can be found at https://github.com/shallowlearner93/NeuroMDAVIS-FS.

## Authors’ contribution

**CM**: Conceptualization, Methodology, Data curation, Data analysis, Formal analysis, Implementation, Writing – Initial Draft Preparation. **VD**: Conceptualization, Methodology, Data curation, Data analysis, Formal analysis, Investigation, Writing – Review & Editing. **DBS**: Conceptualization, Methodology, Data curation, Data analysis, Formal analysis, Investigation, Writing – Review & Editing. **RKD**: Conceptualization, Methodology, Writing – Review & Editing, Overall Supervision.

## Acknowledgements

**RKD** acknowledges the Department of Biotechnology, Government of India, for supporting this research in the form of the grant (Sanction Order Number: BT/PR40176/BTIS/137/84/2023).

## Footnotes

1 https://www.ncdirindia.org/All_Reports/Report_2020/default.aspx

2 https://seer.cancer.gov/statfacts/html/lungb.html

