## Supplementary material for "Multi-Omics Integration for Identification of Prognostic Molecular Signatures for Survival Stratification in Lung Cancer"

February 28, 2026

### Supplementary Material

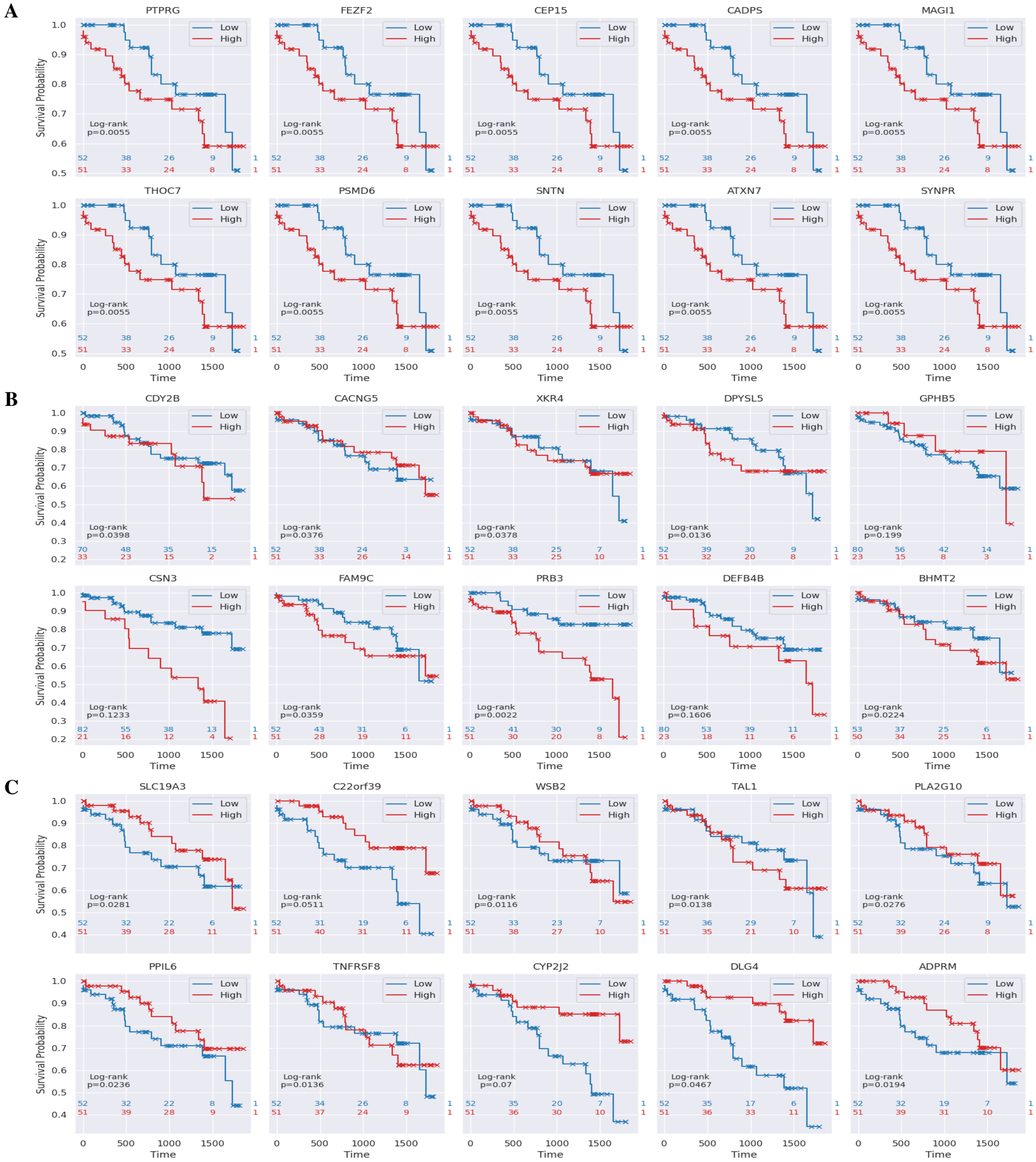


Figure S1: Kaplan-Meier curves based on high and low-expression values of each of the top features selected by NeuroMDAVIS-FS from (A) Protein, (B) RNA, and (C) CNV omics modalities, while applied on the LSCC cohort. Patients were stratified into high and low-expression groups based on the median value of each feature. Log-rank tests were conducted to assess statistical significance in survival differences.

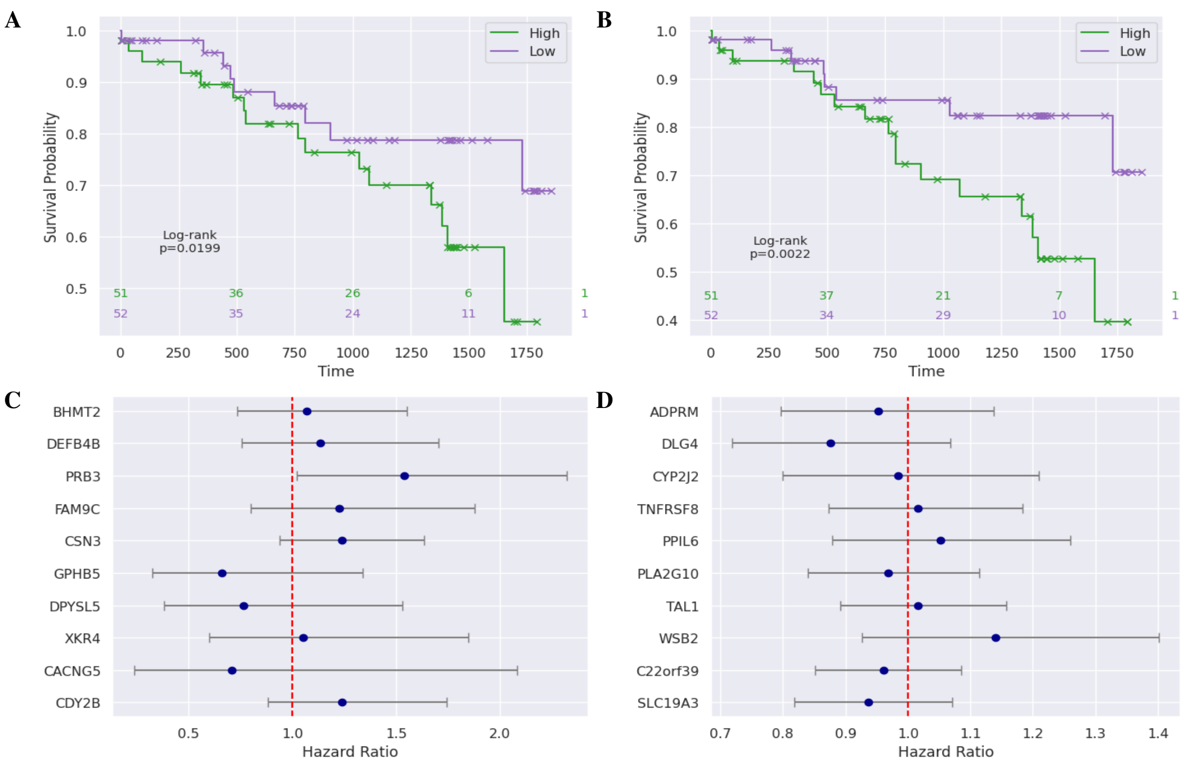


Figure S2: Kaplan-Meier curve based on high and low risk scores obtained from a CoxPH regression applied using (A) top RNAs and (B) top proteins, selected by NeuroMDAVIS-FS. Hazard scores of (C) top RNAs and (D) top proteins, selected by NeuroMDAVIS-FS. These results were obtained utilizing the LSCC cohort only.

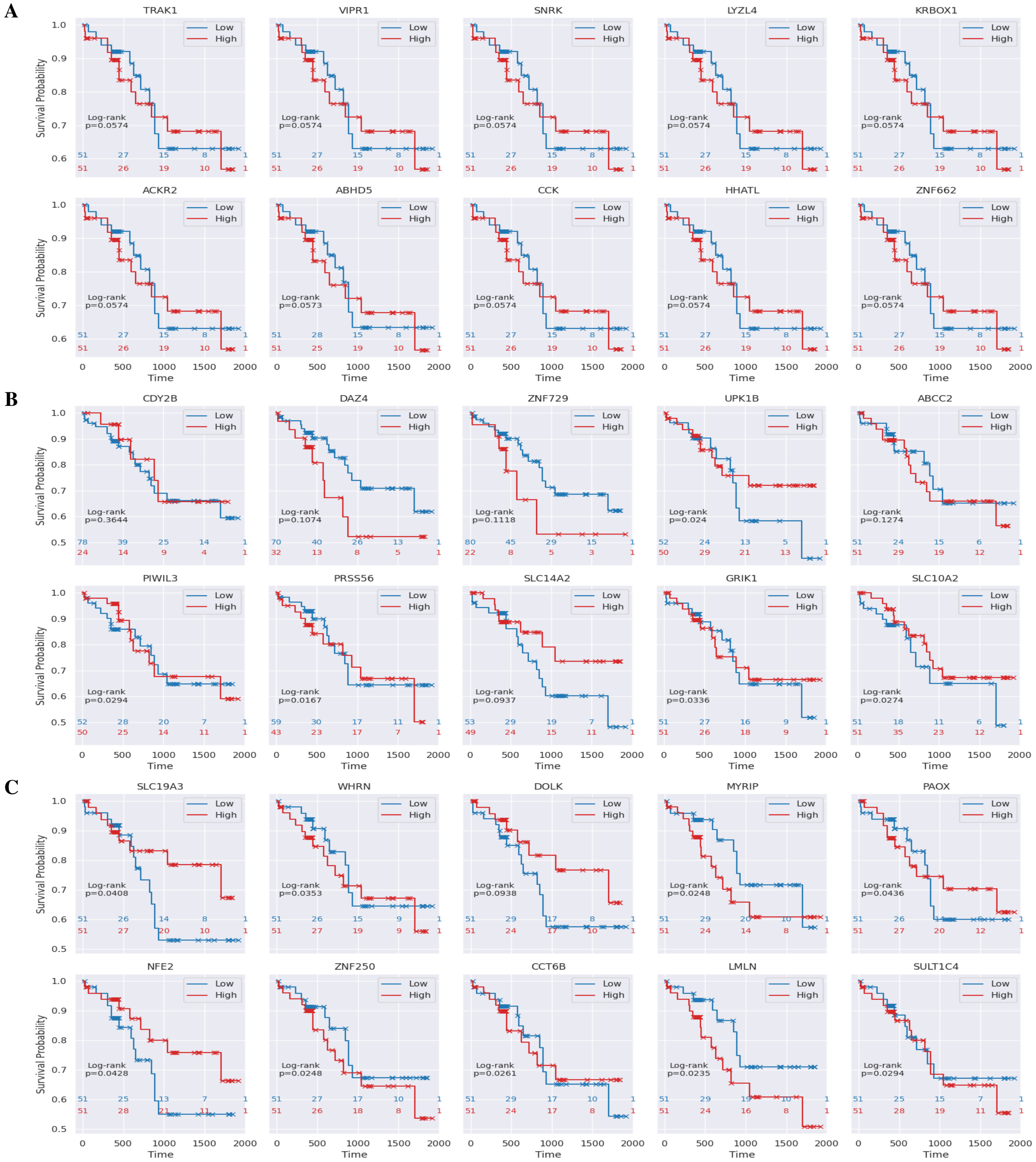


Figure S3: Kaplan-Meier curves based on high and low-expression values of each of the top features selected by NeuroMDAVIS-FS from (A) Protein, (B) RNA, and (C) CNV omics modalities, while applied on the LUAD cohort. Patients were stratified into high and low-expression groups based on the median value of each feature. Log-rank tests were conducted to assess statistical significance in survival differences.

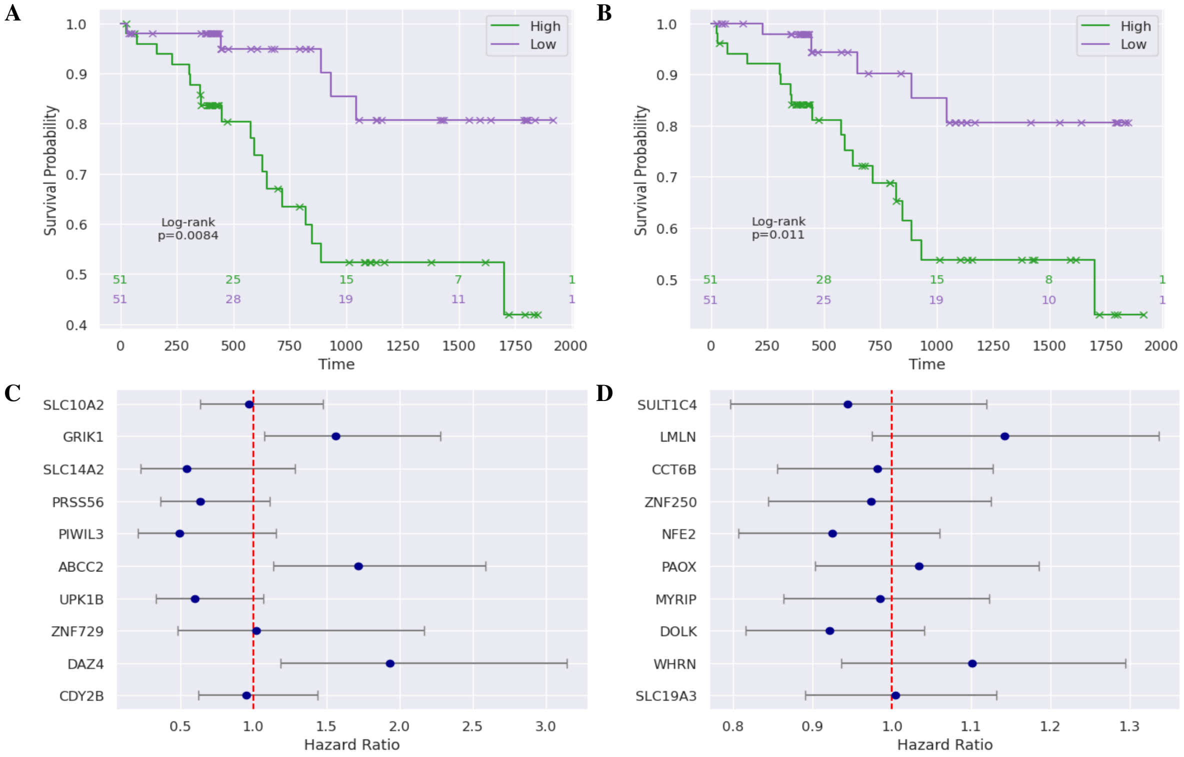


Figure S4: Kaplan-Meier curve based on high and low risk scores obtained from a CoxPH regression applied using (A) top RNAs and (B) top proteins, selected by NeuroMDAVIS-FS. Hazard scores of (C) top RNAs and (D) top proteins, selected by NeuroMDAVIS-FS. These results were obtained utilizing the LUAD cohort only.
